# Genetic analyses of gynecological disease identify genetic relationships between uterine fibroids and endometrial cancer, and a novel endometrial cancer genetic risk region at the *WNT4* 1p36.12 locus

**DOI:** 10.1101/2020.11.09.20228114

**Authors:** Pik Fang Kho, Sally Mortlock, Endometrial Cancer Association Consortium, International Endometriosis Genetics Consortium, Peter A.W. Rogers, Dale R. Nyholt, Grant W. Montgomery, Amanda B. Spurdle, Dylan M. Glubb, Tracy A. O’Mara

**Author notes:** **Corresponding Author** Dr Tracy O’Mara, PhD, Molecular Cancer Epidemiology Group, QIMR Berghofer Medical Research Institute, 300 Herston Road, Brisbane QLD Australia 4006. These authors contributed equally to the work (Dylan M. Glubb, Tracy A. O’Mara). **Declarations**. **Funding** PFK is supported by an Australian Government Research Training Program PhD Scholarship and QIMR Berghofer Postgraduate Top-Up Scholarship, TAO’M, GWM and ABS are supported by NHMRC Investigator Fellowships (APP1173170, GNT1177194 and APP1177524). This work was supported by National Health and Medical Research Council (NHMRC) Project Grants (APP1109286, GNT1026033, GNT1105321 and GNT1147846). Funding sources had no role in study design, data curation and analysis, data interpretation, report writing and submission for publication. **Conflict of interest/Competing interests** The authors declare no potential conflicts of interest. **Ethics approval** This work used summary-level GWAS meta-analysis results, and thus ethical approval was not required. **Consent to participate** Not applicable. **Consent for publication** Not applicable. **Availability of data and material (data transparency)** Summary-level GWAS meta-analysis results for PCOS, uterine fibroids and endometrial cancer that support the findings of this study are available at the NHGRI-EBI GWAS Catalog (https://www.ebi.ac.uk/gwas/downloads/summary-statistics). Other data generated during this study are included in this article and its supplementary information files or are available on reasonable request. **Code availability** Not applicable.

## Abstract

Endometriosis, polycystic ovary syndrome (PCOS) and uterine fibroids have been proposed as endometrial cancer risk factors; however, disentangling their relationships with endometrial cancer is complicated due to shared risk factors and comorbidities. Using genome-wide association study (GWAS) data, we explored the relationships between these non-cancerous gynecological diseases and endometrial cancer risk by assessing genetic correlation, causal relationships and shared risk loci. We found significant genetic correlation between endometrial cancer and PCOS, and uterine fibroids. Adjustment for genetically predicted body mass index (a risk factor for PCOS, uterine fibroids and endometrial cancer) substantially attenuated the genetic correlation between endometrial cancer and PCOS but did not affect the correlation with uterine fibroids. Mendelian randomization analyses provided evidence of a causal relationship between only uterine fibroids and endometrial cancer. Gene-based analyses revealed risk regions shared between endometrial cancer and endometriosis, and uterine fibroids. Multi-trait GWAS analysis of endometrial cancer and the genetically correlated gynecological diseases identified a novel genome-wide significant endometrial cancer risk locus at 1p36.12, which replicated in an independent endometrial cancer dataset. Interrogation of functional genomic data at 1p36.12 revealed biologically relevant genes, including *WNT4* which is necessary for the development of the female reproductive system. In summary, our study provides genetic evidence for a causal relationship between uterine fibroids and endometrial cancer. It further provides evidence that the comorbidity of endometrial cancer, PCOS and uterine fibroids may partly be due to shared genetic architecture. Notably, this shared architecture has revealed a novel genome-wide risk locus for endometrial cancer.

## Introduction

Endometriosis, polycystic ovary syndrome (PCOS) and uterine fibroids are three common non-cancerous gynecological diseases affecting 10-15% (Parasar et al. 2017), 6-9% (Azziz et al. 2011) and 5-69% (Stewart et al. 2017) of women of reproductive age, respectively; however, their prevalence is likely underestimated because of under diagnosis (Agarwal et al. 2019; De La Cruz and Buchanan 2017). Although these non-cancerous gynecological diseases primarily affect premenopausal women and endometrial cancer is largely a postmenopausal malignancy, many risk factors are shared with endometrial cancer (e.g. chronic estrogen exposure, inflammation, insulin resistance and obesity (Harris and Terry 2016; Li et al. 2019; Wise et al. 2016)), suggesting some shared biological relationship.

A number of studies have used observational data to assess associations between the three non-cancerous gynecological diseases and endometrial cancer risk, the findings of which have been heterogeneous (Harris and Terry 2016; Johnatty et al. 2020; Li et al. 2019; Wise et al. 2016). Indeed, the use of observational studies to evaluate these associations could be confounded by: (i) the failure to adequately account for potential confounders that are associated with risk of endometrial cancer and/or gynecological disease e.g. oral contraceptive use; (ii) the reliance of disease status classification on self-reported data which is subject to misclassification bias from asymptomatic undiagnosed cases; (iii) misdiagnosis of early stage endometrial cancer as uterine fibroids due to shared clinical presentation (Wise et al. 2016); (iv) detection bias in cohort studies as a result of an increased surveillance for endometrial cancer among patients with non-cancerous gynecological diseases; and (v) the comorbidity of non-cancerous gynecological diseases (Choi et al. 2017; Johnatty et al. 2020; Matalliotaki et al. 2018; Nagai et al. 2015; Uimari et al. 2011; Wise et al. 2007). Thus, it remains difficult to determine from observational studies the precise nature of the relationships between endometrial cancer and these non-cancerous gynecological diseases.

Genome-wide association study (GWAS) data have demonstrated genetic correlation between endometrial cancer and endometriosis (Masuda et al. 2020; Painter et al. 2018), and uterine fibroids (Masuda et al. 2020), which may partly explain the comorbidities of these diseases; whether these comorbidities are due to causal relationships or shared genetic etiology remains to be explained. In this study, we have used a variety of approaches to analyze GWAS data and elucidate relationships between endometrial cancer and non-cancerous gynecological disease (summarized in **Supplementary Figure 1**). Firstly, we have performed genetic correlation analysis, using the largest currently available datasets to clarify the shared genetic risk between the non-cancerous gynecological diseases and endometrial cancer. As inherited genetic variants are less influenced by confounding inherent in observational studies, we have performed genetic causal inference analyses using gynecological disease-associated variants to investigate causal relationships. It is possible that these diseases may not be genetically correlated or causally related to endometrial cancer but share overlapping genetic risk regions. To assess this possibility, we have performed gene-based association analyses. Lastly, we have performed multi-trait GWAS, leveraging genetic correlation between endometrial cancer and non-cancerous gynecological diseases to discover novel GWAS risk loci.

## Materials and Methods

### GWAS data

GWAS summary data publicly available for PCOS (Day et al. 2018) (https://doi.org/10.17863/CAM.27720) and uterine fibroids (Gallagher et al. 2019) (ftp://ftp.ebi.ac.uk/pub/databases/gwas/summary_statistics/GallagherCS_31649266_GCST009158), and via collaboration for endometriosis (Sapkota et al. 2017), were used for all analyses except Mendelian randomization. For PCOS and uterine fibroids, GWAS summary data from the 23andMe, Inc., cohort had been excluded because of restrictions related to data sharing agreements (Day et al. 2018; Gallagher et al. 2019). For Mendelian randomization analyses, risk estimates and respective standard errors of genome-wide significant variants were accessed from the largest published GWAS for each disease (Day et al. 2018; Gallagher et al. 2019; Rahmioglu et al. 2018). Details of studies and sample sizes used in each analysis are shown in **Supplementary Table 1**. Detailed descriptions of the quality control procedures and GWAS analysis can be found in the corresponding publications.

**Table 1.**
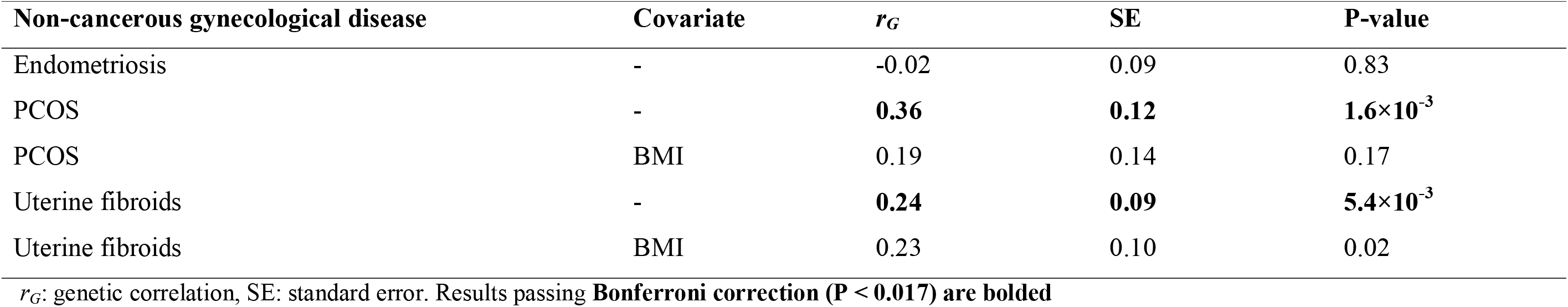
Genetic correlation between non-cancerous gynecological diseases and endometrial cancer.

GWAS summary data for endometrial cancer were available from O’Mara et al. (2018). As the GWAS for endometrial cancer (O’Mara et al. 2018), endometriosis (Rahmioglu et al. 2018), and uterine fibroids (Gallagher et al. 2019) included participants from the UK Biobank (https://www.ukbiobank.ac.uk/), we re-analyzed the endometrial cancer dataset, excluding these participants to avoid sample overlap bias in the two sample Mendelian randomization analysis. This also allowed us to use the UK Biobank endometrial cancer dataset as part of the replication set to confirm multi-trait GWAS results. This revised endometrial cancer GWAS meta-analysis consisted of 12,270 cases and 46,126 controls of European descent. Genetic variants with minor allele frequency (MAF) < 1% and imputation information scorem < 0.4 were excluded, leaving ∼9 million genetic variants. The revised endometrial cancer GWAS data were used only in Mendelian randomization and multi-trait GWAS analyses, while the published endometrial cancer GWAS data (O’Mara et al. 2018) were used in all other analyses. Prior to genetic correlation analysis, genetic variants in the extended human major histocompatibility complex region (26–34 Mb on chromosome 6) were removed due to the complex linkage disequilibrium (LD) structure in this region.

### Genetic correlation between endometrial cancer and non-cancerous gynecological diseases

We used LD Score regression (Bulik-Sullivan et al. 2015) to estimate the genetic correlation between endometrial cancer and each non-cancerous gynecological disease. Genetic correlation analyses were restricted to common HapMap3 variants (MAF > 0.01). To reduce bias from potential residual confounding in genetic correlation analyses, including bias from unknown sample overlap, we used the estimated genetic covariance intercept, obtained without constraint. Genetic correlation values range from -1 to 1; positive values indicated that shared genetic variants have concordant effects across the genome, whereas negative values indicated divergent effects.

Obesity is a major risk factor for endometrial cancer, and is prevalent amongst women with PCOS and uterine fibroids (Ilaria and Marci 2018; Sam 2007). For genetic correlation analysis between endometrial cancer and PCOS or uterine fibroids, we thus additionally corrected for the effect of obesity, as measured by genetically predicted BMI. PCOS and uterine fibroids GWAS were conditioned using summary data from a large GWAS of BMI (Yengo et al. 2018) in GCTA-mtCOJO analysis (Zhu et al. 2018) before performing LD score regression analysis.

### Genetic causal inference tests

We performed two-sample Mendelian randomization analysis to explore potential causal relationships between non-cancerous gynecological diseases and endometrial cancer. Independent (LD *r*^2^ < 0.01) genetic variants associated with the non-cancerous gynecological diseases at genome wide significance (P < 5 × 10^−8^) were used as genetic instruments. The list of genetic instruments and the respective risk association estimates were extracted from the largest GWAS of endometriosis (Rahmioglu et al. 2018), PCOS (Day et al. 2018) and uterine fibroids (Gallagher et al. 2019). We excluded independent genetic variants with ambiguous alleles and intermediate frequencies (i.e., variants with A/T or C/G alleles and minor allele frequency of more than 0.42), leaving 26 variants as genetic instruments for endometriosis, 14 for PCOS and 23 for uterine fibroids.

As the three non-cancerous gynecological diseases mostly affect premenopausal women and endometrial cancer primarily affects postmenopausal women, we performed a unidirectional Mendelian randomization analysis, assessing the effect of genetic predisposition to non-cancerous gynecological disease on endometrial cancer risk. We used inverse variance weighted (IVW) analysis as the primary analysis by regressing the genetic variant-endometrial cancer association on the genetic variant-non-cancerous gynecological disease association, weighted by inverse of their variance. This method has the most power to detect associations although it has a strong assumption of no heterogeneity (potentially resulting from pleiotropy) amongst genetic variants (Hemani et al. 2018); thus, this method assumes all genetic variants for the exposure of interest have a proportional effect on outcome risk.

We also performed several sensitivity analyses for Mendelian randomization that are more robust to heterogeneity amongst genetic variants: MR-Egger, weighted median, and weighted mode analysis. MR-Egger analysis regresses the genetic variant-outcome association on the genetic variant-exposure association, without constraining the regression intercept (Bowden et al. 2015). If the MR-Egger regression intercept is non-zero, it provides evidence that directional horizontal pleiotropy amongst genetic variants is driving the causal estimates (i.e. genetic variation influences the outcome through a pathway other than the exposure and indicates that the ratio of genetic variants with positive and negative pleiotropic effects is not balanced). The MR-Egger regression slope represents a valid effect estimate after adjustment for pleiotropic effects, provided the Instrument Strength Independent of Direct Effect (InSIDE) assumption is met (i.e. the association of a genetic variant with the exposure of interest is independent from its direct effect on outcome) (Bowden et al. 2015). We also performed weighted median (Bowden et al. 2016) and weighted mode (Hartwig et al. 2017) analyses, which are more robust to violation of the InSIDE assumption. Weighted median analysis relies on the assumption that more than 50% of the weights come from valid genetic instruments (Bowden et al. 2016), while weighted mode analysis relies on the assumption that most of the weights come from valid genetic instruments (Hartwig et al. 2017).

Cochran’s Q statistic was used to assess the heterogeneity in the effects of variants (a potential indicator of horizontal pleiotropy) (Bowden et al. 2018), and leave-one-out analysis was used to assess whether a single variant drives the causal association (Hemani et al. 2018).

Two-sample Mendelian randomization analysis was performed using the “TwoSampleMR” (Hemani et al. 2018) package in R. Unless stated otherwise, results with a Bonferroni-corrected p-value for testing the effects of the three non-cancerous gynecological diseases (P < 0.05/3 = 0.017) on endometrial cancer risk were considered statistically significant.

### Gene-based association analysis

To identify genetic risk regions shared between the non-cancerous gynecological diseases and endometrial cancer, we performed gene-based analysis using the fast and flexible set-based association test (fastBAT) (Bakshi et al. 2016). fastBAT was used to perform an enrichment analysis on GWAS risk variants, located within 50kb of gene regions, for the non-cancerous gynecological cancers and endometrial cancer. A random sample of 10,000 unrelated participants from the UK Biobank was used as the reference panel in these analyses. We applied a false discovery rate (FDR) < 0.05 for the gene-based analysis, and adjacent risk-associated genes were considered a single risk region if within 1 Mb of each other.

### Multi-trait analysis of GWAS (MTAG)

MTAG (Turley et al. 2018) was used to improve endometrial cancer risk loci discovery through joint analysis of endometrial cancer and non-cancerous gynecological diseases that showed evidence of genetic correlation with endometrial cancer (i.e. PCOS and uterine fibroids). GWAS summary statistics were used as input and bivariate LD score regression was used to account for sample overlap. Using pre-computed LD scores for Europeans, MTAG analysis was performed on common variants (MAF > 0.01). Alleles of genetic variants were aligned across GWAS, and only variants present across included studies were assessed by MTAG. The final number of included variants for MTAG was 4,734,443. Summary statistics were produced for each trait where effect sizes and standard error estimates could be interpreted as the output from a single-trait GWAS.

### Replication of novel endometrial cancer GWAS risk loci

MTAG assumes that the variance-covariance matrix across traits is homogenous across the genome, but it is likely some variants are null for one trait and not null for another trait(s). Violation of this assumption could increase false positive discovery in MTAG (discussed in (Turley et al. 2018)). To address this issue, we tested the replication of novel genome-wide significant endometrial cancer risk variants from MTAG in an independent GWAS meta-analysis using data from the Finnish Biobank Study (FinnGen; https://www.finngen.fi/en) and the UK Biobank. Endometrial cancer GWAS summary statistics for 566 cases and 75,822 controls were downloaded directly from FinnGen (data freeze 2; http://r2.finngen.fi/). Quality control procedures for the FinnGen GWAS data are described in https://finngen.gitbook.io/documentation/. For UK Biobank, we performed an endometrial cancer GWAS using genotype and phenotype data obtained under the application number 25331. Endometrial cancer cases were defined based on ICD10 code (C54) in the data fields of 40006, 41270 and 41202. Controls were selected randomly from unrelated women participants (π□ < 0.1) with no history of any cancers. GWAS was performed on 1,866 cases and 18,660 controls using REGENIE (Mbatchou et al. 2020) to implement a logistic mixed model, adjusting for genotyping array and the top 10 principal components. A genetic relationship matrix was included in the model as a random effect to account for cryptic relatedness and population stratification. As recommended by REGENIE, we excluded genetic variants with MAF < 0.01, minor allele count below 100, genotype missingness above 10% and variants which deviated from Hardy-Weinberg equilibrium (P value < 1×10^−15^). After quality control exclusion, a total of 9,789,172 SNPs remained in the GWAS analysis.

To create a replication set, the FinnGen and UK Biobank GWAS results were meta-analysed by a fixed-effect inverse variance weighted model using “meta” software in R. Novel endometrial cancer genome-wide significant variants identified by MTAG were considered to have replicated if they had the same effect direction, and a P-value < 0.05 for association in the replication set.

### Identification of candidate target genes at the 1p36.12 endometrial cancer risk locus

We used previously generated promoter-associated HiChIP chromatin looping data from endometrial (one immortalized and three tumor) cell lines (O’Mara et al. 2019) to explore potential regulatory interactions between credible causal risk variants and gene promoters at the 1p36.12 locus. Credible causal risk variants were defined as variants with P-value for association within two orders of magnitude of the lead variant P-value. We also explored the candidate target genes through overlap of credible causal variants with lead cis-eQTLs from GTEx v8 and the Blood eQTL Browser data (Munz et al. 2020).

## Results

We found endometrial cancer to be significantly genetically correlated with PCOS (*r*_*G*_ = 0.36, se = 0.12) and uterine fibroids (*r*_*G*_ = 0.24, se = 0.09) but not with endometriosis (**Table 1**). After adjusting for genetically predicted BMI, the genetic correlation between PCOS and endometrial cancer was no longer statistically significant, indicating that the initial genetic correlation was, at least partly, mediated by genetically predicted BMI (**Table 1**). In contrast, there was no material difference in the genetic correlation between uterine fibroids and endometrial cancer after adjusting for genetically predicted BMI (**Table 1**), consistent with a previous observation of no significant differences in BMI for endometrial cancer cases with or without uterine fibroids (Johnatty et al. 2020).

IVW Mendelian randomization analysis for the effects of genetic predisposition to the non-cancerous gynecological diseases on endometrial cancer provided evidence only for uterine fibroids (**Table 2, Figure 1**). Although sensitivity analyses were not statistically significant, the directionality of the associations between uterine fibroids and endometrial cancer were consistent with the IVW result (**Table 2, Figure 1**). The MR-Egger intercept did not significantly differ from zero (**Table 2**) providing no evidence for confounding by directional horizontal pleiotropy amongst genetic instruments. However, Cochran’s Q statistics indicated evidence of heterogeneity between causal estimates based on individual variants (Cochran’s Q statistics = 42.1, degrees of freedom = 22, P = 6×10^−3^), suggesting that some variants may be associated with endometrial cancer risk through pathways other than uterine fibroids. Leave-one-out analysis showed that no single variant was driving the causal association revealed by the IVW analysis (**Supplementary Figure 2**).

**Table 2.** Genetic causal inference results for effects of non-cancerous gynecological diseases on endometrial cancer.

| Gynecological disease | Genetic causal inference analysis | Beta | SE | P-value |
| --- | --- | --- | --- | --- |
| Endometriosis | IVW | 0.09 | 0.09 | 0.34 |
|  | MR-Egger | 0.44 | 0.30 | 0.15 |
|  | MR-Egger (intercept) | -0.03 | 0.03 | 0.23 |
|  | Weighted median | 0.11 | 0.08 | 0.15 |
|  | Weighted mode | 0.13 | 0.11 | 0.26 |
| PCOS | IVW | -0.05 | 0.04 | 0.26 |
|  | MR-Egger | 0.07 | 0.21 | 0.75 |
|  | MR-Egger (intercept) | -0.02 | 0.03 | 0.57 |
|  | Weighted median | -0.08 | 0.06 | 0.21 |
|  | Weighted mode | -0.19 | 0.12 | 0.13 |
| Uterine fibroids | IVW | **0.17** | **0.07** | **0.01** |
|  | MR-Egger | 0.15 | 0.15 | 0.32 |
|  | MR-Egger (intercept) | 0.00 | 0.01 | 0.91 |
|  | Weighted median | 0.07 | 0.07 | 0.35 |
|  | Weighted mode | 0.01 | 0.12 | 0.96 |

**Figure 1.**
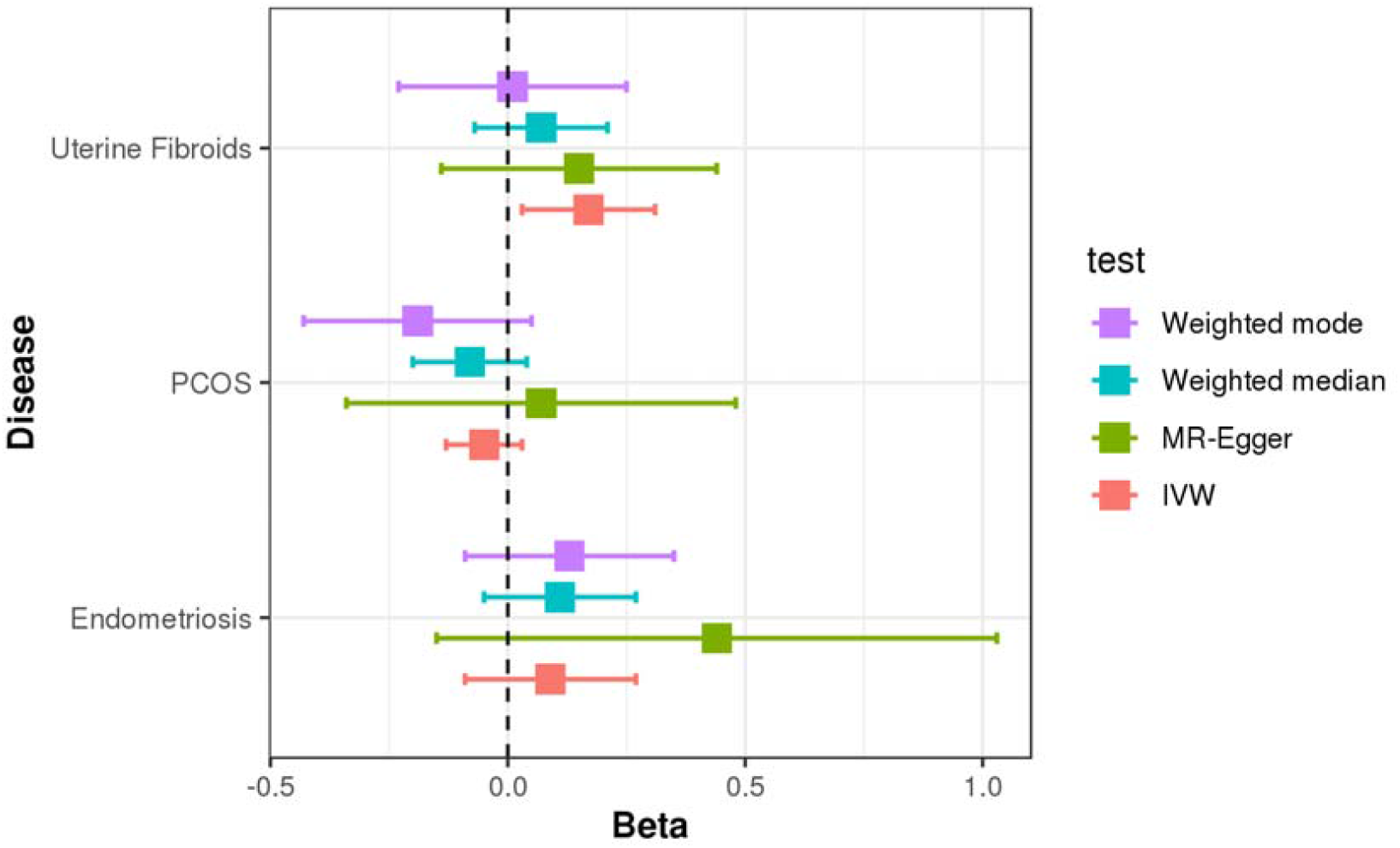
Association between genetic predisposition to non-cancerous gynecological diseases and endometrial cancer, obtained from two-sample Mendelian randomization analysis. The boxes represent the risk of endometrial cancer (beta) per standard deviation increment in genetic predisposition to non-cancerous gynecological disease. Error bars represent 95% confidence intervals.

While genetic correlation analysis assesses the average genetic concordance across the genome for two traits, it does not reveal common genomic regions that harbor trait-associated variation. Further, a lack of evidence for genetic correlation may reflect opposing pleiotropic effects across the genome. Thus, we performed gene-based analyses to identify common risk regions across endometrial cancer and the non-cancerous gynecological diseases. The initial analysis revealed 24 genetic regions associated with endometrial cancer risk, 28 regions with endometriosis risk and 41 regions with uterine fibroids (**Supplementary Table 2**). No associations with PCOS passed FDR < 0.05, potentially reflecting a lack of power due to the small sample size of this cohort. We found four genetic risk regions (3q21.3, 9p21.3, 15q15.1 and 17q21.32), containing seven shared candidate susceptibility genes, were shared between endometriosis and endometrial cancer (**Table 3**). Three of these regions (9p21.3, 15q15.1 and 17q21.32) have independently been associated with the risks of endometrial cancer (O’Mara et al. 2018) and endometriosis through GWAS (Rahmioglu et al. 2018). The LD of lead risk variants at each gene was compared and only one region (17q21.32) demonstrated evidence of a shared genetic risk signal (*r*^2^ > 0.9; **Table 3**). Additionally, we found two genetic risk regions (5p15.33 and 11p13), containing five shared candidate susceptibility genes, were shared between uterine fibroids and endometrial cancer (**Table 3**). 5p15.33 has been associated with uterine fibroids risk through GWAS (Gallagher et al. 2019) while 11p13 has independently associated with uterine fibroids and endometrial cancer risk in GWAS (Gallagher et al. 2019; O’Mara et al. 2018). The LD of lead risk variants at each gene was compared but there was no strong genetic correlation at either 5p15.33 or 11p13 (*r*^2^ ≤ 0.4; **Table 3**), suggesting that the genetic risk signals may be independent.

**Table 3.** Shared candidate endometrial cancer and non-cancerous gynecological diseases risk regions.

| Region | Gene | TopSNP<sub>EC</sub><br>(p-value) | TopSNP<sub>Gyne</sub><br>(p-value) | LD between<br>TopSNPs ($r^2$)* | fastBAT<sub>EC</sub><br>p-value | fastBAT<sub>EC</sub><br>FDR | fastBAT<sub>Gyne</sub><br>p-value | fastBAT<sub>Gyne</sub><br>FDR |
| --- | --- | --- | --- | --- | --- | --- | --- | --- |
| *Endometrial cancer and endometriosis* |  |  |  |  |  |  |  |  |
| 3q21.3 | *RUVBL1* | rs872267<br>($9.95 \times 10^{-7}$) | rs4857864<br>($5.35 \times 10^{-6}$) | 0.47 | $8.49 \times 10^{-6}$ | $8.01 \times 10^{-3}$ | $8.26 \times 10^{-5}$ | 0.043 |
| 9p21.3 | *CDKN2B-AS1* | rs568447<br>($2.68 \times 10^{-6}$) | rs6475610<br>($1.73 \times 10^{-9}$) | $1.0 \times 10^{-4}$ | $7.26 \times 10^{-5}$ | 0.036 | $9.02 \times 10^{-9}$ | $3.55 \times 10^{-5}$ |
| 15q15.1 | *BMF* | rs28371998<br>($6.81 \times 10^{-9}$) | rs7183386<br>($7.23 \times 10^{-6}$) | 0.36 | $1.64 \times 10^{-7}$ | $5.74 \times 10^{-4}$ | $4.93 \times 10^{-6}$ | $9.35 \times 10^{-3}$ |
| 17q21.32 | *CBX1* | rs7225865<br>($1.34 \times 10^{-8}$) | rs10445377<br>($3.20 \times 10^{-7}$) | 0.99 | $3.35 \times 10^{-6}$ | $4.32 \times 10^{-3}$ | $2.63 \times 10^{-5}$ | 0.025 |
| 17q21.32 | *MIR1203* | rs4794505<br>($9.50 \times 10^{-9}$) | rs10445377<br>($3.20 \times 10^{-7}$) | 0.99 | $5.88 \times 10^{-7}$ | $1.44 \times 10^{-3}$ | $4.93 \times 10^{-6}$ | $9.35 \times 10^{-3}$ |
| 17q21.32 | *SKAP1* | rs882380<br>($4.66 \times 10^{-9}$) | rs10445377<br>($3.20 \times 10^{-7}$) | 0.96 | $5.80 \times 10^{-6}$ | $5.93 \times 10^{-3}$ | $7.93 \times 10^{-6}$ | 0.013 |
| 17q21.32 | *SNX11* | rs17681336<br>($1.17 \times 10^{-8}$) | rs10445377<br>($3.20 \times 10^{-7}$) | 0.99 | $1.55 \times 10^{-6}$ | $3.17 \times 10^{-3}$ | $4.51 \times 10^{-6}$ | $9.35 \times 10^{-3}$ |
| *Endometrial cancer and uterine fibroids* |  |  |  |  |  |  |  |  |
| 5p15.33 | *CLPTM1L* | rs2736100<br>($5.17 \times 10^{-6}$) | rs72709458<br>($6.07 \times 10^{-15}$) | 0.28 | $3.93 \times 10^{-7}$ | $1.20 \times 10^{-3}$ | $1.00 \times 10^{-9}$ | $1.53 \times 10^{-6}$ |
| 5p15.33 | *MIR4457* | rs2736100<br>($5.17 \times 10^{-6}$) | rs72709458<br>($6.07 \times 10^{-15}$) | 0.28 | $1.50 \times 10^{-6}$ | $3.17 \times 10^{-3}$ | $8.01 \times 10^{-11}$ | $1.63 \times 10^{-7}$ |
| 5p15.33 | *TERT* | rs2736100<br>($5.17 \times 10^{-6}$) | rs72709458<br>($6.07 \times 10^{-15}$) | 0.28 | $1.72 \times 10^{-6}$ | $3.25 \times 10^{-3}$ | $1.50 \times 10^{-11}$ | $4.06 \times 10^{-8}$ |
| 11p13 | *WT1* | rs10835920<br>($1.33 \times 10^{-8}$) | rs11031731<br>($2.04 \times 10^{-21}$) | 0.20 | $1.86 \times 10^{-5}$ | 0.015 | $2.64 \times 10^{-16}$ | $3.22 \times 10^{-12}$ |
| 11p13 | *WT1-AS* | rs10835920<br>($1.33 \times 10^{-8}$) | rs11031762<br>($9.95 \times 10^{-14}$) | 0.40 | $1.91 \times 10^{-6}$ | $3.34 \times 10^{-3}$ | $1.04 \times 10^{-12}$ | $4.24 \times 10^{-9}$ |

TopSNP: lead variant for gene from fastBAT analysis; EC: endometrial cancer; Gyne: non-cancerous gynecological diseases; LD: linkage disequilibrium; FDR: false discovery rate.
\*LD was estimated using the EUR 1000Genomes reference panel.

Incorporation of the two gynecological diseases genetically correlated with endometrial cancer (uterine fibroids and PCOS) in MTAG revealed ten genome-wide significant risk loci for endometrial cancer (**Table 4, Figure 2**). We observed an inflation of median test statistics in the MTAG result (λ = 1.06), which was likely due to a polygenic signal (LD score regression intercept = 0.98, se = 0.01) rather than population stratification. Two of the risk loci (5p15.33 and 1p36.12) were novel endometrial cancer genome-wide risk loci. We assessed both these risk loci in an independent endometrial cancer dataset and found that only the association at the 1p36.12 locus replicated (Table 4).

**Table 4.**
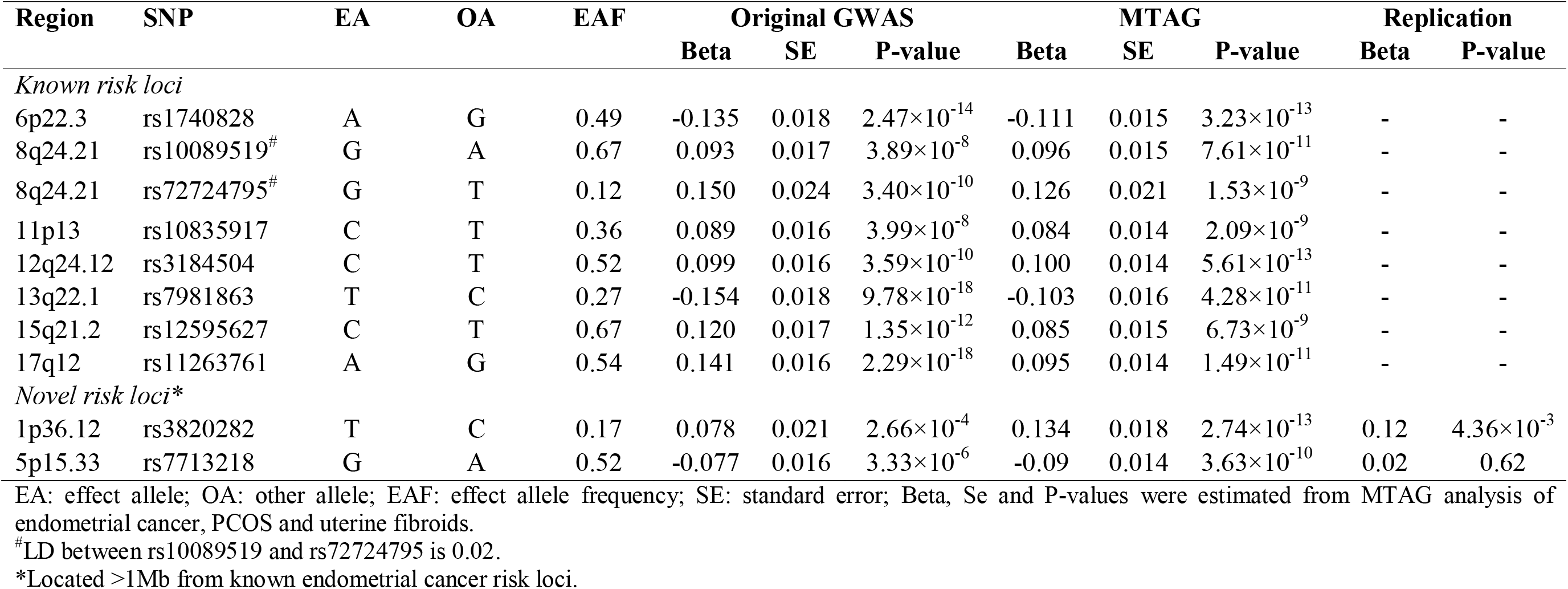
Genome-wide significant endometrial cancer risk loci identified using MTAG.

**Figure 2.**
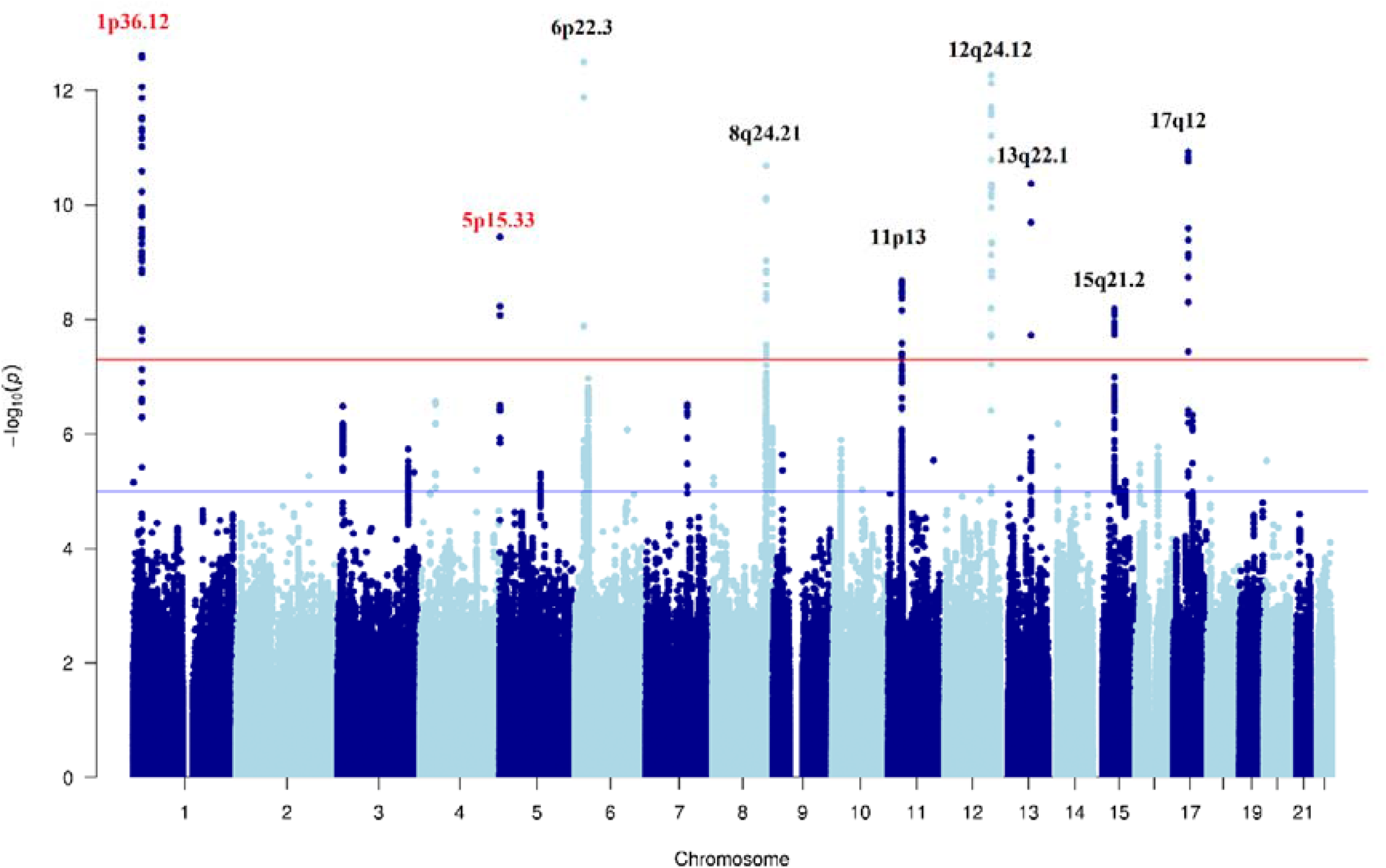
Manhattan plot of MTAG result for endometrial cancer risk. Known endometrial cancer GWAS risk loci are marked in black, and novel genome-wide significant risk loci that are located >1Mb from known endometrial cancer risk loci in red.

To identify candidate target genes at the replicated novel endometrial cancer GWAS risk locus (1p36.12), we intersected candidate causal variants with promoter-associated chromatin loops from four endometrial (immortalized and tumor) cell lines (O’Mara et al. 2019). We identified six candidate target genes through chromatin looping, including *WNT4* for which a candidate causal risk variant was revealed as a lead eQTL in lung tissue (**Supplementary Tables 3 and 4; Figure 3**). Additionally, we identified *CDC42* as a candidate target gene through a candidate causal risk variant located in a chromatin looping anchor at its promoter (**Supplementary Table 3; Figure 3**). Furthermore, candidate causal risk variants were lead eQTLs for *CDC42* in peripheral blood (Westra et al. 2013) (**Supplementary Table 4)**, providing additional evidence for regulatory targeting.

**Figure 3.**
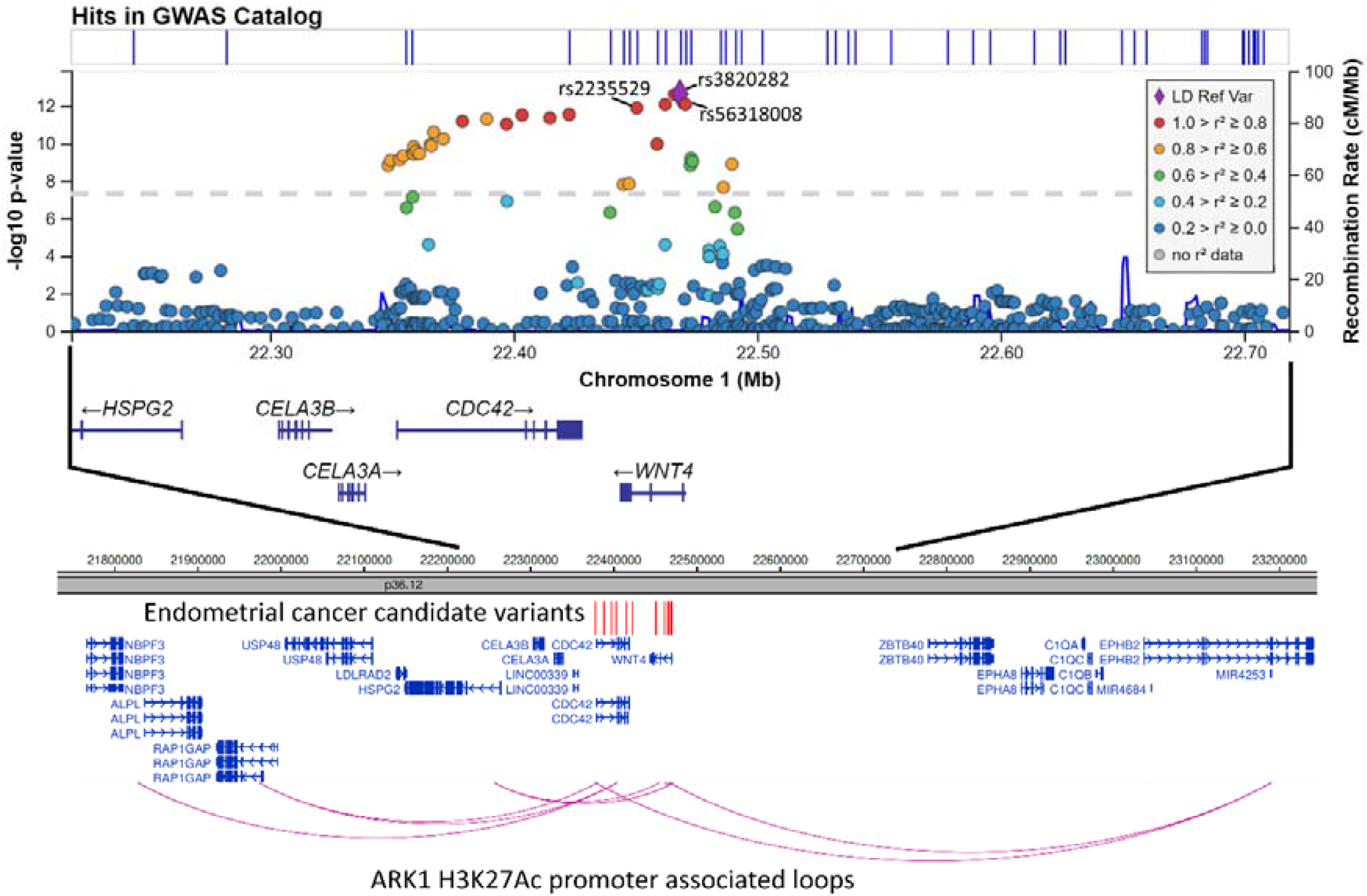
The upper panel depicts a regional association plot for the 1p36.12 novel endometrial cancer risk locus. Genetic variants at the locus are plotted by their genomic position (hg19) and MTAG -log_10_(P) for association with endometrial cancer risk is on the left y-axis. Recombination rate (cM/Mb) is on the right y-axis and plotted as blue lines. The color of the circles indicates the level of linkage disequilibrium between each variant and the lead variant, rs3820282 (purple diamond), from the 1000 Genomes 2014 EUR reference panel (see legend, inset). The lower panel shows promoter-associated chromatin looping at 1p36.12 identified from HiChIP analysis of the ARK-1 endometrial cancer cell line. Promoter-associated loops that intersect with candidate causal variants (shown as red vertical lines) are shown as purple arcs.

## Discussion

Using large-scale genome-wide datasets, we observed evidence of positive genetic correlation between endometrial cancer and PCOS, and uterine fibroids, but not endometriosis. The observed genetic correlation between endometrial cancer and PCOS was at least partly mediated by genetically predicted BMI, consistent with the role of BMI as a risk factor for both PCOS and endometrial cancer. Mendelian randomization analysis provided evidence for a causal relationship only between genetic predisposition to uterine fibroids and endometrial cancer risk. Gene-based analyses revealed several genetic risk regions shared between endometrial cancer and endometriosis, and uterine fibroids. This included one apparent joint genetic risk signal, for endometrial cancer and endometriosis at 17q21.32. Multi-trait GWAS analysis, including endometrial cancer and the genetically correlated gynecological diseases identified two novel genome-wide significant risk loci for endometrial cancer, one of which (1p36.12) replicated in an independent endometrial cancer dataset. Lastly, functional analyses highlighted *CDC42* and *WNT4* as candidate target genes at the 1p36.12 endometrial cancer risk locus.

Two previous studies have reported a positive genetic correlation between endometriosis and endometrial cancer (Masuda et al. 2020; Painter et al. 2018), but we found no evidence for such genetic correlation. This discrepancy may be related to: i) the smaller sample sets used by the prior studies; ii) the ethnicity studied (Masuda et al. (2020) analyzed a Japanese population); or iii) the different genetic correlation analysis approaches used. For example, unlike Painter et al. (2018), we used an unconstrained LD score regression intercept to account for potential residual confounding, resulting in a conservative estimate of genetic correlation. Indeed, we found the estimated genetic covariance intercept to be significantly different from zero, suggesting the presence of bias from population stratification and/or sample overlap. The null results from the genetic causal inference analyses of endometriosis and endometrial cancer are concordant with observational studies that observed no associations after controlling for ascertainment bias by excluding recent endometriosis diagnosis (Melin et al. 2007; Olson et al. 2002; Rowlands et al. 2011).

Although there was minimal evidence for genetic correlation or a causal relationship between endometriosis and endometrial cancer, we identified four shared genetic risk regions, three of which (9p21.3, 15q15.1 and 17q21.32) have independently been associated with risk of both diseases through GWAS (O’Mara et al. 2018; Rahmioglu et al. 2018). Notably, the shared candidate susceptibility genes at 9p21.3 (C*DKN2B-AS1*), 15q15.1 (*BMF*) and 17q21.32 (*CBX1, MIR1203, SKAP1* and *SNX11)* have been previously identified as candidate target genes at endometrial cancer GWAS risk loci through promoter-associated chromatin looping studies (O’Mara et al. 2019). The remaining shared endometriosis and endometrial cancer risk region at 3q21.3 has not been independently identified by GWAS for either disease and may represent a novel GWAS risk locus for both in future studies. Indeed, this region was recently reported as an endometrial cancer risk region in a cross-cancer GWAS meta-analysis of endometrial, breast, ovarian and prostate cancer (Kar et al. 2020).

We found PCOS and endometrial cancer to be genetically correlated but no association was observed in genetic causal inference analyses, concordant with observational studies that account for the effect of obesity (Fearnley et al. 2010; Zucchetto et al. 2009). These findings are consistent with our observation of substantial attenuation in genetic correlation between PCOS and endometrial cancer after adjusting for genetic components of BMI.

We detected evidence of positive genetic correlation between uterine fibroids and endometrial cancer risk, consistent with observational studies (Fortuny et al. 2009; Rowlands et al. 2011; Wise et al. 2016). IVW Mendelian randomization analysis provided evidence of a causal relationship between genetic predisposition to uterine fibroids and endometrial cancer risk. However, Cochran’s Q statistics showed evidence that variants used in the IVW analysis had heterogeneous effects, suggesting that not all variants that increase uterine fibroids risk are expected to increase endometrial cancer risk. Although results from subsequent sensitivity analyses that are robust to the presence of varying levels of pleiotropy were not statistically significant, they showed concordant effect directions with IVW result. It is important to note that the Mendelian randomization sensitivity analyses have lower power to detect causal relationships compared with IVW analysis. As genetic causal inference tests rely on the statistical power of GWAS used, future larger GWAS are required to provide more accurate causal estimates and thus greater confidence with regards to the nature of the relationship between uterine fibroids and endometrial cancer.

Gene-based analysis revealed two genetic risk regions (5p15.33 and 11p13) that were shared by endometrial cancer and uterine fibroids. The 11p13 shared risk region has been associated with the risks of uterine fibroids (Gallagher et al. 2019) and endometrial cancer (O’Mara et al. 2018) in GWAS. From the gene-based analysis, we identified *WT1* and *WT1-AS* as candidate susceptibility genes for both uterine fibroids and endometrial cancer at 11p13. Consistent with this finding, we had previously established both genes as candidate targets of endometrial cancer risk GWAS variation through promoter-associated chromatin looping studies (O’Mara et al. 2019) and *WT1* had also been identified through chromatin looping as a candidate target of uterine fibroids risk variants (Rafnar et al. 2018). *WT1* encodes a transcription factor that is essential for urogenital development (reviewed by Roberts (2005)) and in the GTEx database of tissue gene expression it is most highly expressed in the uterus (https://gtexportal.org/home/). These observations suggest that alteration of uterine *WT1* expression by endometrial cancer and uterine fibroids genetic risk variation may affect susceptibility to these diseases.

The 5p15.33 region was found to associate with endometrial cancer risk through both the gene-based analysis and the multi-trait GWAS. However, the multi-trait GWAS association did not replicate in the independent endometrial cancer dataset, with discordant effect directions and non-overlapping confidence intervals. Previously, this region has associated with uterine fibroids risk in a GWAS (Gallagher et al. 2019), with endometrial cancer risk in a candidate locus study (Carvajal-Carmona et al. 2015) and in a cross-cancer GWAS meta-analysis of endometrial cancer and ovarian cancer (Glubb et al. 2021). The gene-based analysis at this region revealed three candidate risk genes that were shared between uterine fibroids and endometrial cancer. The most biologically relevant of these genes is *TERT*, which encodes telomerase reverse transcriptase and maintains chromosomal stability by elongating the telomere (Rubtsova et al. 2012). Relevantly, chromosomes in uterine fibroids (Bonatz et al. 1998; Rogalla et al. 1995) and in endometrial tumors (reviewed by Alnafakh et al. (2019)) have been shown to have shorter telomere length. Indeed, a recent Mendelian randomization study found genetically predicted telomere length to be strongly associated with endometrial cancer risk (Telomeres Mendelian Randomization et al. 2017).

The novel 1p36.12 endometrial cancer risk locus, revealed by the multi-trait GWAS, replicated in the independent endometrial cancer GWAS dataset. Genetic variation at this region has associated with traits that are genetically correlated or causally related to endometrial cancer (e.g. heel bone mineral density (Morris et al. 2019), body mass index (Pulit et al. 2019), diabetes (Vujkovic et al. 2020), age at menarche (Kichaev et al. 2019) and ovarian cancer (Kuchenbaecker et al. 2015)). Furthermore, genetic variation at 1p36.12 has associated with endometriosis and the lead endometrial cancer risk variant from the multi-trait GWAS also represents a GWAS risk signal for pelvic organ prolapse (Olafsdottir et al. 2020). Promoter-associated chromatin looping data from endometrial cell lines highlighted seven candidate target genes, two of which (*CDC42* and *WNT4*) were supported by candidate causal risk variants that represent eQTLs for these genes. The lead candidate causal risk variant at 1p36.12 (rs3820282) and two candidate causal variants (rs61768001 & rs12037376) have previously been associated with expression of *CDC42* in blood and a long non-coding RNA (*LINC00339*) in blood and the endometrium (Mortlock et al. 2020). Semi-quantitative chromatin looping analysis in an endometrial cancer cell line demonstrated evidence of an interaction between a region containing rs3820282 and a ∼15 kb region containing the promoter of *LINC00339* (Powell et al. 2016). However, the quantitative chromatin looping data from the HiChIP analysis of the normal immortalized and tumoral endometrial cell lines (O’Mara et al. 2019)), which also has much greater resolution (Lareau and Aryee 2018), did not provide evidence for a physical interaction between *LINC00339* and candidate causal endometrial cancer risk variants.

*LINC00339, CDC42* and *WNT4* all have biological function relevant to endometrial cancer. *LINC00339* has been found to promote oncogenesis in several different cancer types (Gao et al. 2020; Ye et al. 2020; Zhao et al. 2020), although not specifically endometrial cancer. *CDC42* encodes a small GTPase of the Rho-subfamily that regulates cell cycle, cell-cell adhesion, cell migration and cancer progression (Qadir et al. 2015). Notably, CDC42 binds to PAK6 (encoded by an endometrial cancer GWAS risk candidate target gene (O’Mara et al. 2019)) and this complex, which localizes to cell-cell adhesions, is correlated with epithelial colony escape (Morse et al. 2016). *WNT4* encodes a protein that activates WNT/β-catenin signaling and appears to be crucial for the development of the female reproductive system, including the uterus (reviewed in (Biason-Lauber and Konrad 2008)). Moreover, genes belonging to the WNT/β-catenin pathway are frequently mutated in cancer, including the gene encoding β-catenin which is mutated in ∼25% of endometrial tumors (Kandoth et al. 2013). As with *CDC42*, there are also links between *WNT4* and other genes located at endometrial cancer GWAS risk loci, such as *WT1* and *RSPO1* (O’Mara et al. 2018). For example, in the ovary there is evidence of *WNT4* regulation by proteins encoded by both of these genes (Biason-Lauber 2012; Gao et al. 2014) and RSPO protein activity potentiates WNT signaling (Bugter et al. 2021). There also appears to be some connection between *CDC42* and *WNT4*: both genes have been found to be differentially expressed in the endometrium during the menstrual cycle (Powell et al. 2016).

To reduce confounding inherent in the comorbidity observational studies of endometrial cancer and gynecological disease, prospective studies with long follow-up, large sample sizes and case identification using surgical confirmation would ideally be performed. Nevertheless, our study has demonstrated the utility of genetic causal inference analysis as a cost-effective alternative approach for unravelling relationships while reducing bias from unmeasured confounding. However, a limitation of our study is that the sample size of PCOS GWAS (the largest publicly available) was relatively small, reducing power to identify shared genetic risk regions or a causal relationship between PCOS and endometrial cancer. Consequently, these analyses should be revisited when more genome-wide significant variants are revealed in future PCOS GWAS.

In conclusion, our study has provided insights into the comorbidity of non-cancerous gynecological diseases and endometrial cancer by revealing shared genetic risk architecture, a potential causal relationship between uterine fibroids and endometrial cancer, and shared candidate risk regions and genes. Furthermore, our study has leveraged this shared genetic architecture to identify a novel risk locus for endometrial cancer, uncovering biologically relevant candidate target genes and furthering our understanding of endometrial cancer etiology.

## Data Availability

Summary-level GWAS meta-analysis results for PCOS, uterine fibroids and endometrial cancer that support the findings of this study are available at the NHGRI-EBI GWAS Catalog (https://www.ebi.ac.uk/gwas/downloads/summary-statistics). Other data generated during this study are included in this article and its supplementary information files or are available on reasonable request.

## Acknowledgements

This work was conducted using the UK Biobank Resource (application number 25331). We thank the research participants and employees of 23andMe for making this work possible. We thank the participants and investigators of FinnGen study. We thank the many individuals who participated in the Endometrial Cancer Association Consortium and the International Endometriosis Genetics Consortium studies, and the numerous institutions and their staff who supported recruitment. A full list of consortium members and acknowledgements can be found in the Supplementary Note.

## Supplementary Figure

**Supplementary Figure 1.**
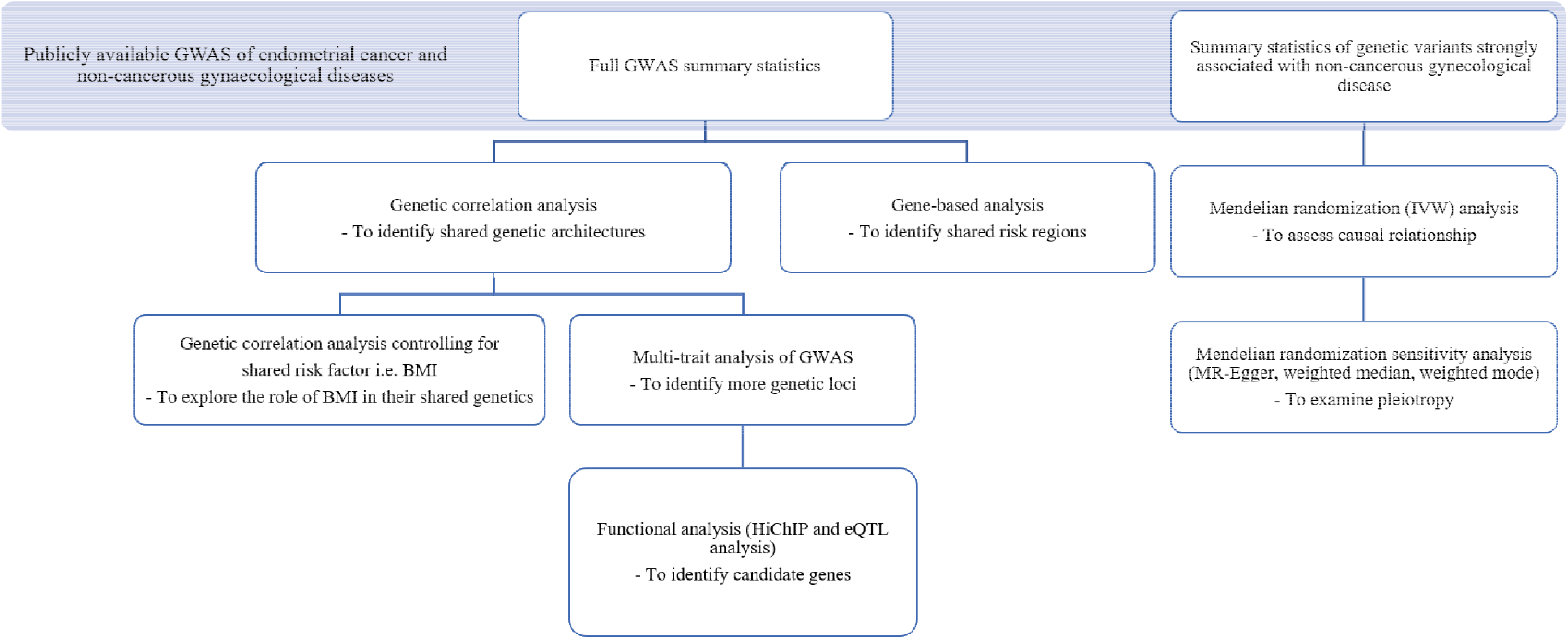
Study design to explore the relationships between non-cancerous gynecological diseases and endometrial cancer. Genetic analyses including genome-wide genetic analysis, gene-based analysis, Mendelian randomization analysis, multi-trait analysis of GWAS and GWAS functional analysis were used in this study.

**Supplementary Figure 2.**
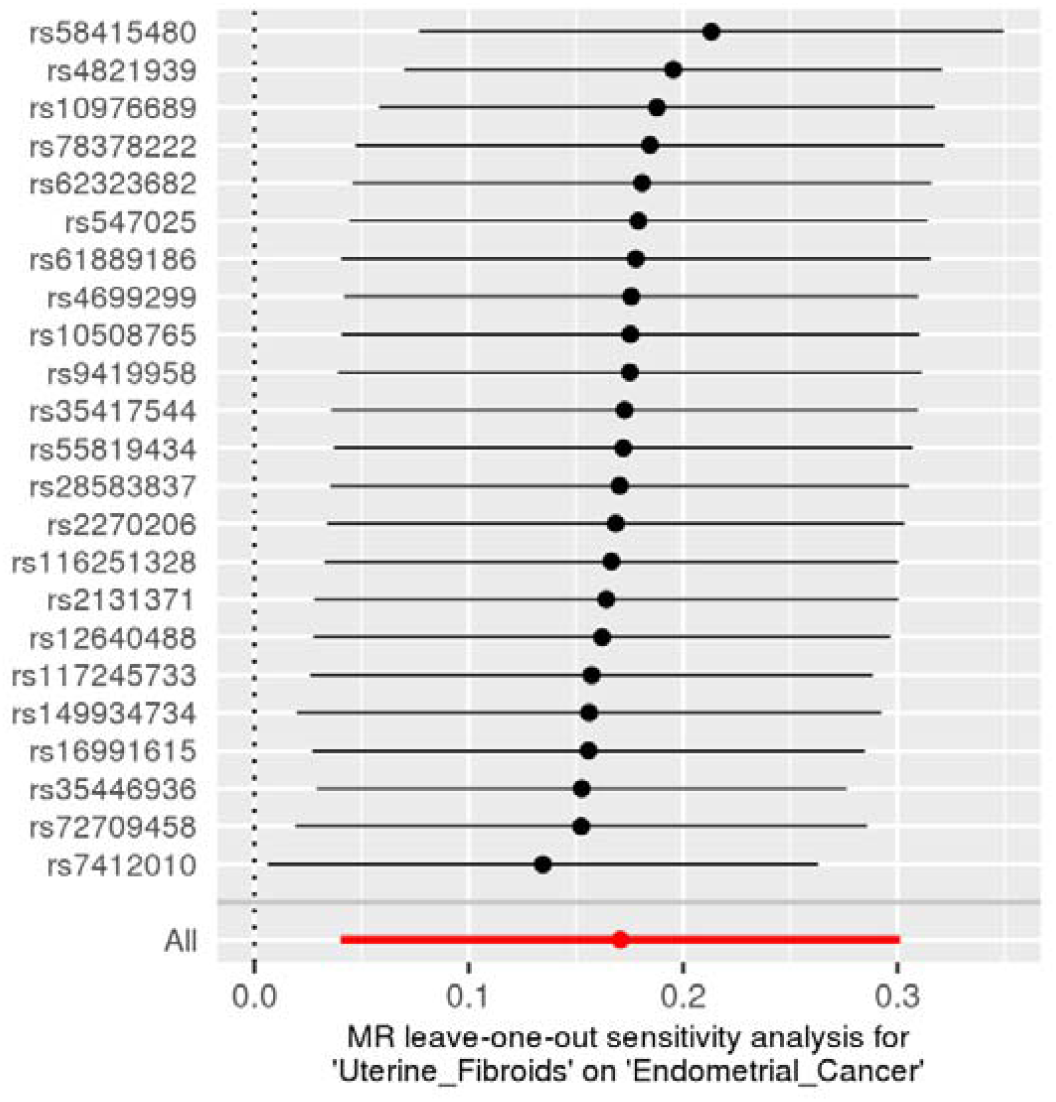
Leave-one-out sensitivity analysis plot for association between genetic predisposition to uterine fibroids and endometrial cancer. Each black dot in the forest plot represents the IVW estimates after excluding the corresponding variant. The plot highlighted in red represents the IVW estimate for all variants.

## Notes

### Competing Interest Statement

The authors have declared no competing interest.

### Author Declarations

This work used summary-level GWAS meta-analysis results, and thus ethical approval was not required.

### Summary of Updates

An error in effect alleles was noticed in the uterine fibroids GWAS which affected Mendelian randomization analyses. This has been updated in this manuscript, significantly altering the results. We have also included multi-trait analysis of GWAS (MTAG) to this version of the manuscript.

