## Supplementary Note for "Genetic analyses of gynecological disease identify genetic relationships between uterine fibroids and endometrial cancer, and a novel endometrial cancer genetic risk region at the *WNT4* 1p36.12 locus"

**Endometrial Cancer Association Consortium Collaborators**

Frederic Amant^1^, Daniela Annibali^1^, Katie Ashton^2-4^, John Attia^2, 5^, Paul L. Auer^6, 7^, Matthias W. Beckmann^8^, Amanda Black^9^, Louise Brinton^9^, Daniel D. Buchanan^10-13^, Stephen J. Chanock^14^, Chu Chen^15^, Maxine M. Chen^16^, Timothy H.T. Cheng^17^, Linda S. Cook^18, 19^, Marta Crous-Bous^16, 20^, Kamila Czene^21^, Immaculata De Vivo^16, 20^, Joe Dennis^22^, Thilo Dörk^23^, Sean C. Dowdy^24^, Alison M. Dunning^25^, Matthias Dürst^26^, Douglas F. Easton^22, 25^, Arif B. Ekici^27^, Peter A. Fasching^8, 28^, Brooke L. Fridley^29^, Christine M. Friedenreich^19^, Montserrat García-Closas^14^, Mia M. Gaudet^30^, Graham G. Giles^11, 31, 32^, Dylan M. Glubb^33^, Ellen L. Goode^34^, Christopher A. Haiman^35^, Per Hall^21, 36^, Susan E. Hankinson^20, 37^, Catherine S. Healey^25^, Alexander Hein^8^, Peter Hillemanns^23^, Shirley Hodgson^38^, Erling Hoivik^39, 40^, Elizabeth G. Holliday^2, 5^, David J. Hunter^16, 41^, Angela Jones^17^, Peter Kraft^16, 42^, Camilla Krakstad^39, 40^, Diether Lambrechts^43, 44^, Loic Le Marchand^45^, Xiaolin Liang^46^, Annika Lindblom^47, 48^, Jolanta Lissowska^49^, Jirong Long^50^, Lingeng Lu^51^, Anthony M. Magliocco^52^, Lynn Martin^53^, Mark McEvoy^5^, Roger L. Milne^11, 31, 32^, Miriam Mints^54^, Rami Nassir^55^, Tracy A. O'Mara^33^, Irene Orlow^46^, Geoffrey Otton^56^, Claire Palles^17^, Paul D.P. Pharoah^22, 25^, Loreall Pooler^35^, Tony Proietto^56^, Timothy R. Rebbeck^57, 58^, Stefan P. Renner^59^, Harvey A. Risch^51^, Matthias Rübner^59^, Ingo Runnebaum^26^, Carlotta Sacerdote^60, 61^, Gloria E. Sarto^62^, Fredrick Schumacher^63^, Rodney J. Scott^2, 4, 64^, V. Wendy Setiawan^35^, Mitul Shah^25^, Xin Sheng^35^, Xiao-Ou Shu^50^, Melissa C. Southey^10, 31, 32^, Amanda B. Spurdle^33^, Emma Tham^47, 65^, Deborah J. Thompson^22^, Ian Tomlinson^17, 53^, Jone Trovik^39, 40^, Constance Turman^16^, David Van Den Berg^35^, Zhaoming Wang^9^, Penelope M. Webb^66^, Nicolas Wentzensen^9^, Stacey J. Winham^67^, Lucy Xia^35^, Yong-Bing Xiang^68^, Hannah P. Yang^9^, Herbert Yu^45^, Wei Zheng^50^

^1^ Department of Obstetrics and Gynecology, Division of Gynecologic Oncology, University Hospitals KU Leuven, University of Leuven, Leuven, Belgium. ^2^ Hunter Medical Research Institute, John Hunter Hospital, Newcastle, New South Wales, Australia. ^3^ Centre for Information Based Medicine, University of Newcastle, Callaghan, New South Wales, Australia.
^4^ Discipline of Medical Genetics, School of Biomedical Sciences and Pharmacy, Faculty of Health, University of Newcastle, Callaghan, New South Wales, Australia. ^5^ Centre for Clinical Epidemiology and Biostatistics, School of Medicine and Public Health, University of Newcastle, Callaghan, New South Wales, Australia. ^6^ Cancer Prevention Program, Fred Hutchinson Cancer Research Center, Seattle, WA, USA. ^7^ Zilber School of Public Health, University of Wisconsin-Milwaukee, Milwaukee, WI, USA. ^8^ Department of Gynecology and Obstetrics, Comprehensive Cancer Center ER-EMN, University Hospital Erlangen, Friedrich-Alexander-University Erlangen-Nuremberg, Erlangen, Germany. ^9^ Division of Cancer Epidemiology and Genetics, National Cancer Institute, Bethesda, MD, USA. ^10^ Department of Clinical Pathology, The University of Melbourne, Melbourne, Victoria, Australia. ^11^ Centre for Epidemiology and Biostatistics, Melbourne School of Population and Global Health, The University of Melbourne, Melbourne, Victoria, Australia. ^12^ Genomic Medicine and Family Cancer Clinic, Royal Melbourne Hospital, Parkville, Victoria, Australia. ^13^ University of Melbourne Centre for Cancer Research, Victorian Comprehensive Cancer Centre, Parkville, Victoria, Australia. ^14^ Division of Cancer Epidemiology and Genetics, National Cancer Institute, National Institutes of Health, Department of Health and Human Services, Bethesda, MD, USA. ^15^ Epidemiology Program, Fred Hutchinson Cancer Research Center, Seattle, WA, USA. ^16^ Department of Epidemiology, Harvard T.H. Chan School of Public Health, Boston, MA, USA. ^17^ Wellcome Trust Centre for Human Genetics and Oxford NIHR Biomedical Research Centre, University of Oxford, Oxford, UK. ^18^ University of New Mexico Health Sciences Center, University of New Mexico, Albuquerque, NM, USA. ^19^ Department of Cancer Epidemiology and Prevention Research, Alberta Health Services, Calgary, AB, Canada. ^20^ Channing Division of Network Medicine, Department of Medicine, Brigham and Women's Hospital and Harvard Medical School, Boston, MA, USA. ^21^ Department of Medical Epidemiology and Biostatistics, Karolinska Institutet, Stockholm, Sweden. ^22^ Centre for Cancer Genetic Epidemiology, Department of Public Health and Primary Care, University of Cambridge, Cambridge, UK. ^23^ Gynaecology Research Unit, Hannover Medical School, Hannover, Germany. ^24^ Department of Obstetrics and Gynecology, Division of Gynecologic Oncology, Mayo Clinic, Rochester, MN, USA. ^25^ Centre for Cancer Genetic Epidemiology, Department of Oncology, University of Cambridge, Cambridge, UK.
^26^ Department of Gynaecology, Jena University Hospital - Friedrich Schiller University, Jena, Germany. ^27^ Institute of Human Genetics, University Hospital Erlangen, Friedrich-Alexander University Erlangen-Nuremberg, Comprehensive Cancer Center Erlangen-EMN, Erlangen, Germany. ^28^ David Geffen School of Medicine, Department of Medicine Division of Hematology and Oncology, University of California at Los Angeles, Los Angeles, CA, USA.
^29^ Department of Biostatistics, Kansas University Medical Center, Kansas City, KS, USA.
^30^ Department of Population Science, American Cancer Society, Atlanta, GA, USA. ^31^ Cancer Epidemiology Division, Cancer Council Victoria, Melbourne, Victoria, Australia. ^32^ Precision Medicine, School of Clinical Sciences at Monash Health, Monash University, Clayton, Victoria, Australia. ^33^ Department of Genetics and Computational Biology, QIMR Berghofer Medical Research Institute, Brisbane, Queensland, Australia. ^34^ Department of Health Science Research, Division of Epidemiology, Mayo Clinic, Rochester, MN, USA. ^35^ Department of Preventive Medicine, Keck School of Medicine, University of Southern California, Los Angeles, CA, USA. ^36^ Department of Oncology, Södersjukhuset, Stockholm, Sweden. ^37^ Department of Biostatistics & Epidemiology, University of Massachusetts, Amherst, Amherst, MA, USA. ^38^ Department of Clinical Genetics, St George's, University of London, London, UK. ^39^ Centre for Cancer Biomarkers CCBIO, Department of Clinical Science, University of Bergen, Bergen, Norway.
^40^ Department of Obstetrics and Gynecology, Haukeland University Hospital, Bergen, Norway. ^41^ Nuffield Department of Population Health, University of Oxford, Oxford, UK. ^42^ Program in Genetic Epidemiology and Statistical Genetics, Harvard T.H. Chan School of Public Health, Boston, MA, USA. ^43^ VIB Center for Cancer Biology, Leuven, Belgium. ^44^ Laboratory for Translational Genetics, Department of Human Genetics, University of Leuven, Leuven, Belgium. ^45^ Epidemiology Program, University of Hawaii Cancer Center, Honolulu, HI, USA.
^46^ Department of Epidemiology and Biostatistics, Memorial Sloan-Kettering Cancer Center, New York, NY, USA. ^47^ Department of Molecular Medicine and Surgery, Karolinska Institutet, Stockholm, Sweden. ^48^ Department of Clinical Genetics, Karolinska University Hospital, Stockholm, Sweden. ^49^ Department of Cancer Epidemiology and Prevention, M. Sklodowska-Curie Cancer Center, Oncology Institute, Warsaw, Poland. ^50^ Division of Epidemiology, Department of Medicine, Vanderbilt Epidemiology Center, Vanderbilt-Ingram Cancer Center, Vanderbilt University School of Medicine, Nashville, TN, USA. ^51^ Chronic Disease Epidemiology, Yale School of Public Health, New Haven, CT, USA. ^52^ Department of Anatomic Pathology, Moffitt Cancer Center & Research Institute, Tampa, FL, USA. ^53^ Institute of Cancer and Genomic Sciences, University of Birmingham, Birmingham, UK. ^54^ Department of Women's and Children's Health, Karolinska Institutet, Stockholm, Sweden. ^55^ Department of Biochemistry and Molecular Medicine, University of California Davis, Davis, CA, USA. ^56^ School of Medicine and Public Health, University of Newcastle, Callaghan, New South Wales, Australia. ^57^ Harvard T.H. Chan School of Public Health, Boston, MA, USA. ^58^ Dana-Farber Cancer Institute, Boston, MA, USA. ^59^ Department of Gynaecology and Obstetrics, University Hospital Erlangen, Friedrich-Alexander University Erlangen-Nuremberg, Comprehensive Cancer Center Erlangen-EMN, Erlangen, Germany. ^60^ Center for Cancer Prevention (CPO-Peimonte), Turin, Italy. ^61^ Human Genetics Foundation (HuGeF), Turino, Italy. ^62^ Department of Obstetrics and Gynecology, School of Medicine and Public Health, University of Wisconsin, Madison, WI, USA. ^63^ Department of Population and Quantitative Health Sciences, Case Western Reserve University, Cleveland, OH, USA. ^64^ Division of Molecular Medicine, Pathology North, John Hunter Hospital, Newcastle, New South Wales, Australia. ^65^ Clinical Genetics, Karolinska Institutet, Stockholm, Sweden. ^66^ Population Health Department, QIMR Berghofer Medical Research Institute, Brisbane, Queensland, Australia. ^67^ Department of Health Sciences Research, Division of Biomedical Statistics and Informatics, Mayo Clinic, Rochester, MN, USA. ^68^ State Key Laboratory of Oncogene and Related Genes & Department of Epidemiology, Shanghai Cancer Institute, Renji Hospital, Shanghai Jiaotong University School of Medicine, Shanghai, China.

**International Endometriosis Genetics Consortium Collaborators**

Yadav Sapkota^1,2^, Valgerdur Steinthorsdottir^3^, Andrew P. Morris^4,5^, Amelie Fassbender^6,7^, Nilufer Rahmioglu^5^, Immaculata De Vivo^8,9^, Julie E. Buring^8,10^, Futao Zhang^11^, Todd L. Edwards^12^, Sarah Jones^13^, Dorien O^6,7^, Daniëlle Peterse^6,7^, Kathryn M. Rexrode^8,10^, Paul M. Ridker^8,10^, Andrew J. Schork^14,15^, Stuart MacGregor^1^, Nicholas G. Martin^1^, Christian M. Becker^16^, Sosuke Adachi^17^, Kosuke Yoshihara^17^, Takayuki Enomoto^17^, Atsushi Takahashi^18^, Yoichiro Kamatani^18^, Koichi Matsuda^19^, Michiaki Kubo^18^, Gudmar Thorleifsson^3^, Reynir T. Geirsson^20,21^, Unnur Thorsteinsdottir^3,21^, Leanne M. Wallace^1,11^, iPSYCH-SSI-Broad Groupw, Jian Yang^11^, Digna R. Velez Edwards^22^, Mette Nyegaard^23,24^, Siew-Kee Low^18^, Krina T. Zondervan^5,16^, Stacey A. Missmer^8,9^, Thomas D’Hooghe^6,7,25^, Grant W. Montgomery^1,11^, Daniel I. Chasman^8,10^, Kari Stefansson^3,21^, Joyce Y. Tung^26^ & Dale R. Nyholt^1,27^.

^1^Department of Genetics and Computational Biology, QIMR Berghofer Medical Research Institute, Brisbane, Queensland 4006, Australia. ^2^Department of Epidemiology and Cancer Control, St. Jude Children’s Research Hospital, Memphis, Tennessee 38105, USA. ^3^ deCODE Genetics/Amgen, 101 Reykjavik, Iceland. ^4^Department of Biostatistics, University of Liverpool, Liverpool L69 3GL, UK. ^5^Wellcome Trust Centre for Human Genetics, University of Oxford, Oxford OX3 7BN, UK. ^6^ KULeuven, Department of Development and Regeneration, Organ systems, 3000 Leuven, Belgium. ^7^Department of Obstetrics and Gynaecology, Leuven University Fertility Centre, University Hospital Leuven, 3000 Leuven, Belgium. ^8^Harvard T.H. Chan School of Public Health, Boston, Massachusetts 02115, USA. ^9^ Channing Division of Network Medicine, Department of Medicine, Brigham and Women’s Hospital and Harvard Medical School, Boston, Massachusetts 02115, USA. ^10^Division of Preventive Medicine, Brigham and Women’s Hospital, Boston, Massachusetts 02215, USA. ^11^ Institute for Molecular Bioscience, The University of Queensland, Brisbane, Queensland 4072, Australia. ^12^ Institute of Medicine and Public Health, Vanderbilt University Medical Center, Nashville, Tennessee 37203, USA. ^13^ Vanderbilt Genetics Institute, Division of Epidemiology, Institute of Medicine and Public Health, Department of Medicine, Vanderbilt University Medical Center, Nashville, Tennessee 37203, USA. ^14^ Cognitive Science Department, University of California, San Diego, La Jolla, California 92093, USA. ^15^ Institute of Biological Psychiatry, Mental Health Centre Sct. Hans, Copenhagen University Hospital, DK-2100 Copenhagen, Denmark. ^16^ Endometriosis CaRe Centre, Nuffield Dept of Obstetrics & Gynaecology, University of Oxford, John Radcliffe Hospital, Oxford OX3 9DU, UK. ^17^Department of Obstetrics and Gynecology, Niigata University Graduate School of Medical and Dental Sciences, Niigata 950-2181, Japan. ^18^ Center for Integrative Medical Sciences, RIKEN, Yokohama 230-0045, Japan. ^19^ Institute of Medical Sciences, The University of Tokyo, Tokyo 108-8639, Japan. ^20^Department of Obstetrics and Gynecology, Landspitali University Hospital, 101 Reykjavik, Iceland. ^21^ Faculty of Medicine, School of Health Sciences, University of Iceland, 101 Reykjavik, Iceland. ^22^Vanderbilt Genetics Institute, Vanderbilt Epidemiology Center, Institute of Medicine and Public Health, Department of Obstetrics and Gynecology, Vanderbilt University Medical Center, Nashville, Tennessee 37203, USA. ^23^Department of Biomedicine, Aarhus University, DK8000 Aarhus, Denmark. ^24^ iPSYCH, The Lundbeck Foundation Initiative for Integrative Psychiatric Research, DK-2100 Copenhagen, Denmark. ^25^Global Medical Affairs Fertility, Research and Development, Merck KGaA, Darmstadt, Germany. ^26^ 23andMe, Inc., 899 W. Evelyn Avenue, Mountain View, California 94041, USA. ^27^ Institute of Health and Biomedical Innovation, Queensland University of Technology, Queensland 4059, Australia.

**Endometrial Cancer Association Consortium Acknowledgements and Funding**

We thank the participants in the endometrial cancer studies included in our study. The Endometrial Cancer Association Consortium genome-wide association analyses were supported by the National Health and Medical Research Council of Australia (APP552402, APP1031333, APP1109286, APP1111246 and APP1061779), the U.S. National Institutes of Health (R01-CA134958), European Research Council (EU FP7 Grant), Wellcome Trust Centre for Human Genetics (090532/Z/09Z) and Cancer Research UK. OncoArray genotyping of ECAC cases was performed with the generous assistance of the Ovarian Cancer Association Consortium (OCAC), which was funded through grants from the U.S. National Institutes of Health (CA1X01HG007491-01 (C.I. Amos), U19-CA148112 (T.A. Sellers), R01-CA149429 (C.M. Phelan) and R01-CA058598 (M.T. Goodman); Canadian Institutes of Health Research (MOP-86727 (L.E. Kelemen)) and the Ovarian Cancer Research Fund (A. Berchuck). We particularly thank the efforts of Cathy Phelan. OncoArray genotyping of the BCAC controls was funded by Genome Canada Grant GPH-129344, NIH Grant U19 CA148065, and Cancer UK Grant C1287/A16563. All studies and funders are listed in O’Mara et al (2018).

**International Endometriosis Genetics Consortium Acknowledgements and Funding**

We acknowledge all the study participants in 11 individual endometriosis studies that provided an opportunity for the current study. We also thank many hospital directors and staff, gynaecologists, general practitioners and pathology services in Australia who provided assistance with confirmation of diagnoses. We thank the research participants and employees of 23andMe for making this work possible. We thank the subjects of the Icelandic deCODE study for their participation. We thank research staff and clinicians for providing diagnostic confirmation for the OX data set. We would like to express our gratitude to the staff and members of the Biobank Japan and Laboratory for Statistical Analysis, RIKEN Center for Integrative Medical Sciences for their outstanding assistance. The QIMR study was supported by grants from the National Health and Medical Research Council (NHMRC) of Australia (241,944, 339,462, 389,927, 389,875, 389,891, 389,892, 389,938, 443,036, 442,915, 442,981, 496,610, 496,739, 552,485, 552,498, 1,026,033 and 1,050,208), the Cooperative Research Centre for Discovery of Genes for Common Human Diseases (CRC), Cerylid Biosciences (Melbourne) and donations from N. Hawkins and S. Hawkins. Analyses of the QIMRHCS and OX GWAS were supported by the Wellcome Trust (WT084766/Z/08/Z) and makes use of WTCCC2 control data generated by the Wellcome Trust Case-Control Consortium (awards 076113 and 085475). The iPSYCH study was funded by The Lundbeck Foundation, Denmark (R102-A9118, R155-2014-1724 ), and the research has been conducted using the Danish National Biobank resource supported by the Novo Nordisk Foundation. A full list of the investigators who contributed to the generation of these data is available from http://www.wtccc.org.uk. D.R.N. was supported in part by the NHMRC Fellowship (613674) and ARC Future Fellowship (FT0991022) schemes. E.G.H. (631096) and G.W.M. (339446, 619667) were supported by the NHMRC Fellowships Scheme. S.M. is supported by an Australian Research Council Future Fellowship. A.P.M. was supported by a Wellcome Trust Senior Research Fellowship (award WT098017). N.R. was supported by funding from the Medical Research Council UK (MR/K011480/1). This study was funded by the BioBank Japan project, which is supported by the Ministry of Education, Culture, Sports, Sciences and Technology of Japanese government.
